# Serological evidence supports the transmission of clade Ib mpox virus by professional sex workers and spread within households in South Kivu, DRC

**DOI:** 10.1101/2025.03.27.25324753

**Authors:** Leandre Murhula Masirika, Luca M. Zaeck, Pacifique Ndishimye, Jean Claude Udahemuka, Saria Otani, Frank M. Aarestrup, Leonard Schuele, Babs E. Verstrepen, Corine H. GeurtsvanKessel, Jean Pierre Musabyimana, Justin Bengehya Mbiribindi, Jules Minega Ndoli, Bas B. Oude Munnink, Freddy Siangoli Belesi, Marion P.G. Koopmans, Rory D. de Vries

**Author notes:** corresponding author: Leandre Murhula Masirika and Rory D. de Vries. First authors contributed equally (Leandre Murhula Masirika, Luca M. Zaeck). Last authors contributed equally (Marion P.G. Koopmans, Rory D. de Vries).

## Abstract

Understanding secondary attack rates is a key knowledge gap in the ongoing clade Ib mpox virus (MPXV) outbreak in the Democratic Republic of the Congo. Here, we report the first serological study to investigate the extent of local MPXV clade Ib transmission, performed in South Kivu, DRC. Sera were collected in November and December 2023 (n=120), and May 2024 (n=48) from professional sex workers (PSW) and visitors of 25 bars with reports of mpox cases. In 18% and 17% of these sera, respectively, serological evidence for MPXV infection was detected, indicating that PSW played an important role in MPXV clade Ib transmission in this region. Additionally, sera from 108 direct contacts of mpox cases from 34 households were collected between September 2023 and May 2024. At least one serum sample tested seropositive in 50% of households. Serological studies are needed to comprehend the extent and severity of the ongoing MPXV outbreak, and may be used to guide targeted vaccination strategies, particularly for high-risk groups.

## Main text

The ongoing mpox outbreaks in DRC, followed by the spread of mpox virus (MPXV) clade Ib from South Kivu, DRC, to neighboring countries, were reasons to call for coordinated actions to stop MPXV spread.^1-3^ To this end, the World Health Organization (WHO) declared a Public Health Emergency of International Concern (PHEIC) in August 2024.^4^ The Africa Centers for Disease Control and Prevention (Africa CDC), in collaboration with the WHO, developed a continental response plan that describes the steps needed to combat mpox.^5^ This included the need for developing laboratory capacity, public health capacity, and vaccines. A pledge for vaccines enabled a first round of vaccination with modified vaccinia Ankara – Bavarian Nordic (MVA-BN) in DRC, initially targeting healthcare workers and risk groups.^6,7^ To target vaccination to specific risk groups to interrupt further transmission, it is important to understand the extent of MPXV circulation. This is difficult to ascertain as the focal point of the outbreak is in regions with little health system capacity.

MPXV clade Ib was first detected in Kamituga, South Kivu, DRC, in September 2023,^8^ where it was associated with a high exposure risk through professional sex workers (PSW).^9,10^ Rapid person-to-person transmission appears to have driven the outbreak, including cross-border transmissions.^11^ As the causative virus is distinct from MPXV historically found in the region,^8^ there are questions regarding the traits of MPXV clade Ib. Initial case descriptions suggest that the associated disease is less severe than disease caused by MPXV clade Ia, but the pattern of spread suggests increased rates of human-to-human transmission, corroborated by an enrichment of APOBEC3-type mutations indicative thereof.^3,9,12^ Better understanding of secondary attack rates is a key knowledge gap. Serological studies can help to understand both the size of the disease pyramid and the true extent of MPXV clade Ib spread. These serological studies are also the cornerstone for understanding and evaluating the immunogenicity of the MVA-BN vaccine, currently being administered in the region to individuals with or without prior exposures through infection and/or vaccination.

Since the spread of MPXV clade Ib and the declaration of the PHEIC, serological studies in Central Africa have been lacking.^2^ Here, we describe a serological study among individuals exposed to an index case in bars with PSW or households, and a pre-outbreak control cohort. Sera from the exposed cohort were collected after the beginning of the clade Ib outbreak in South Kivu, DRC, between September 2023 and May 2024. For the control cohort, archived serum samples were used, which were collected at least two years prior to the ongoing MPXV clade Ib outbreak in an area that has not reported mpox before. These studies were performed to (1) determine the level of MPXV exposure in PSW, (2) determine the level of exposure through non-sexual household transmission, and (3) assess the possibility to perform serological assays at local laboratories.

Sera were obtained from a total of 276 potentially exposed individuals in DRC. Sera collected from 51 individuals pre-outbreak served as negative controls. Most of the sera from the exposed cohort were obtained from individuals working in or visiting bars (n=168), collected either in November and December 2023 (n=120), or May 2024 (n=48) (**Table 1**). These individuals include bar personnel and mining workers, but most sera were collected from PSW (n=113). Additionally, sera were obtained from household contacts of confirmed mpox cases (n=108) between September 2023 and May 2024 (**Table 1**). Out of the 276 sera obtained from these exposed exposure cohorts, 21 (7.6%) were human immunodeficiency virus-1 (HIV-1) seropositive. According to the UNAIDS, the national seroprevalence in the general population of DRC was 0.7% in 2023; however, in PSW specifically, the reported HIV seroprevalence was 7.5%, similar to what we find in this cohort.^13^

**Table 1.**
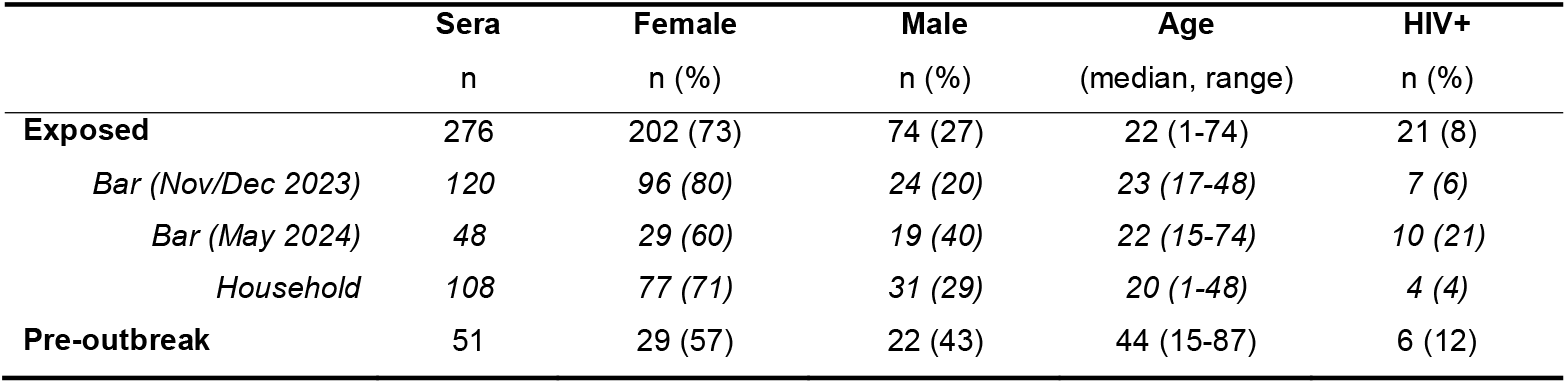
Cohort characteristics. Characteristics for exposed (n=276) and pre-outbreak (n=51) individuals. Sera from exposed individuals were categorized as obtained from individuals in bars or household contacts. Three samples from the bar group collected in Nov/Dec 2023 (exposed cohort) and one sample from the pre-outbreak cohort could not be tested for HIV seropositivity due to insufficient sample volumes.

|  | Sera | Female | Male | Age | HIV+ |
| --- | --- | --- | --- | --- | --- |
|  | n | n (%) | n (%) | (median, range) | n (%) |
| **Exposed** | 276 | 202 (73) | 74 (27) | 22 (1-74) | 21 (8) |
| *Bar (Nov/Dec 2023)* | 120 | 96 (80) | 24 (20) | 23 (17-48) | 7 (6) |
| *Bar (May 2024)* | 48 | 29 (60) | 19 (40) | 22 (15-74) | 10 (21) |
| *Household* | 108 | 77 (71) | 31 (29) | 20 (1-48) | 4 (4) |
| **Pre-outbreak** | 51 | 29 (57) | 22 (43) | 44 (15-87) | 6 (12) |

As an initial screening, we measured levels of vaccinia virus (VACV)-binding antibodies in all sera by ELISA, as described previously,^14^ to assess potential orthopoxvirus exposure. Among all potentially exposed individuals, 62 out of 276 (23%) had detectable VACV-binding antibodies (**Table 2**). In sera collected from bars in November and December 2023, or May 2024, VACV-binding antibodies were measured in 25/120 (21%) and 10/48 (21%) samples, respectively. In sera collected from households, the ELISA-positivity rate was 27/108 (25%) (**Figure 1A**). Since most of the samples in the potentially exposed cohort were obtained from individuals younger than 44 years (267 out of 276, 97%), and smallpox vaccination in DRC stopped around 1980,^15^ most of the individuals were expected to be serologically naive for VACV-binding antibodies. Therefore, the high seropositivity rate was likely caused by MPXV infection, although exposure to other (animal) pox viruses cannot be ruled out. Antibody levels were comparable among male and female individuals, and across age groups (**Figure S1A**). VACV-binding antibodies were detected in 6 out of 51 (12%) of the pre-outbreak controls (**Figure 1A**). Four of those individuals were in the age range where they could have received historic smallpox vaccination.

**Table 2.**
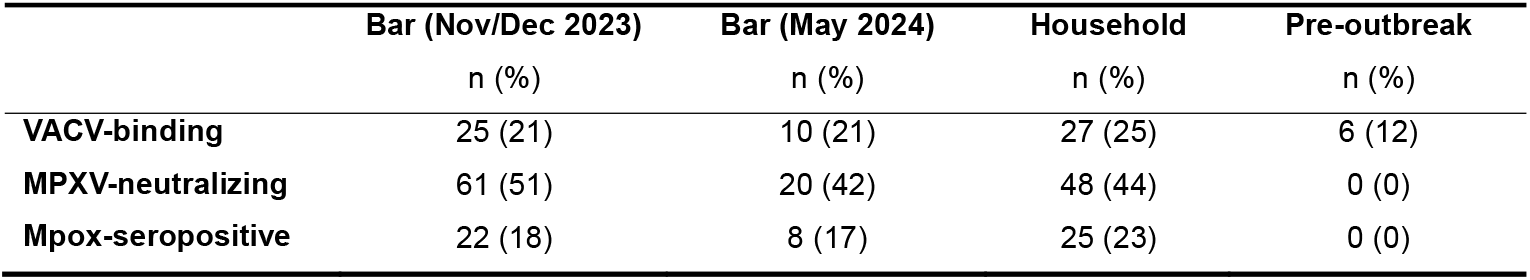
Detection of VACV-binding and MPXV-neutralizing antibodies and associated mpox seropositivity. Mpox seropositivity is defined as a detectable titer in a VACV-binding antibody ELISA (>10) and confirmation by MPXV-neutralizing antibody titers threefold over background (>35).

|  | Bar (Nov/Dec 2023) | Bar (May 2024) | Household | Pre-outbreak |
| --- | --- | --- | --- | --- |
|  | n (%) | n (%) | n (%) | n (%) |
| **VACV-binding** | 25 (21) | 10 (21) | 27 (25) | 6 (12) |
| **MPXV-neutralizing** | 61 (51) | 20 (42) | 48 (44) | 0 (0) |
| **Mpox-seropositive** | 22 (18) | 8 (17) | 25 (23) | 0 (0) |

**Figure 1.**
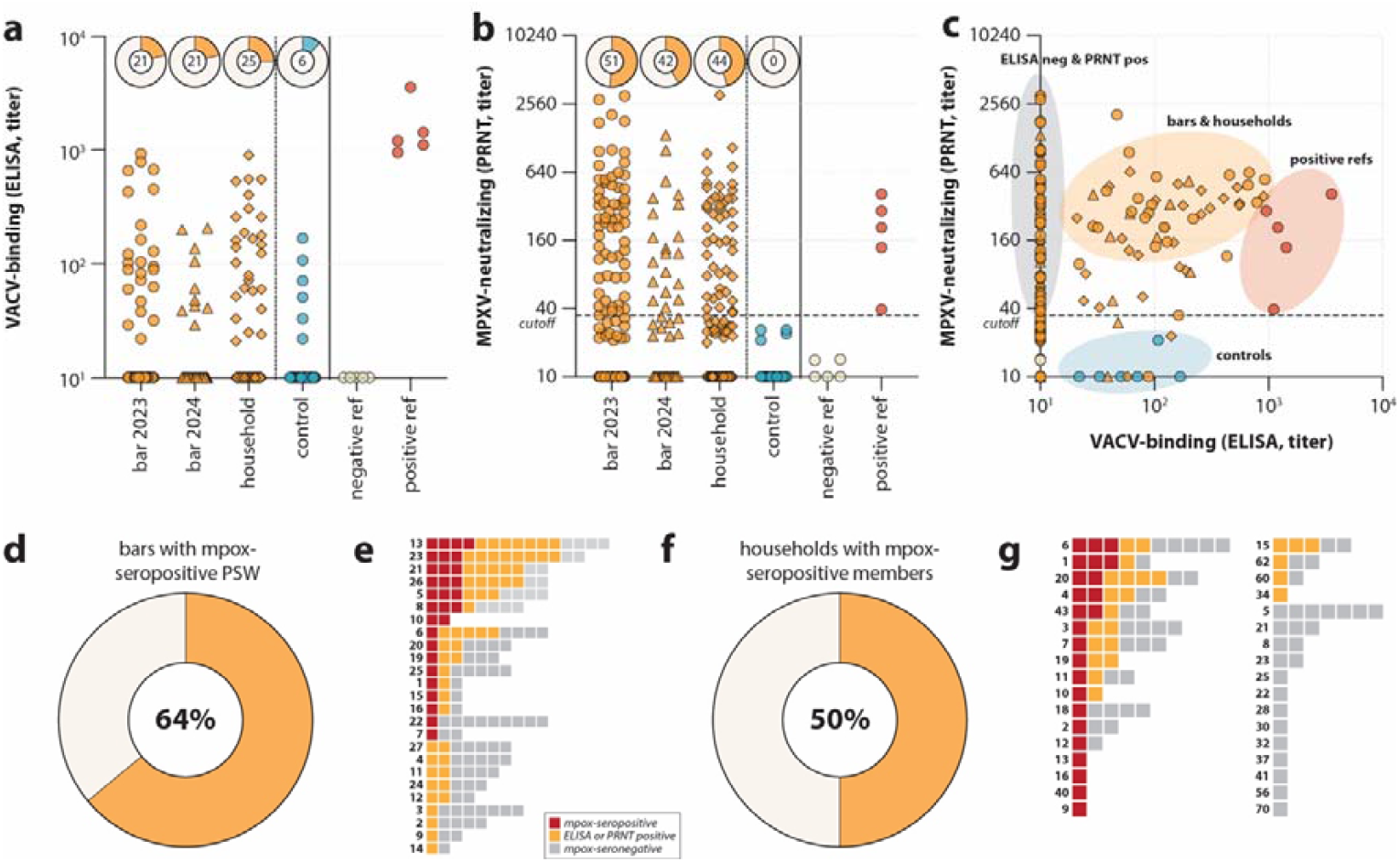
VACV-binding and MPXV-neutralizing antibodies in sera from exposed (high-risk) and unexposed (pre-outbreak) individuals in DRC. (A) VACV-binding antibodies in sera obtained from bars in 2023, bars in 2024, households (all exposed, orange), controls (unexposed, teal), and validation sera (grey and red). Fraction of seropositive sera is indicated per group. (B) MPXV-neutralizing antibodies in sera shown in A. (C) Correlation between VACV-binding binding and MPXV-neutralizing antibodies in exposed individuals (orange), controls (teal), and validation sera from vaccinated individuals (red). Colored bubbles indicate manually defined clusters. (D) Percentage of bars with seropositive professional sex workers (PSW). (E) Serostatus from all individuals in bars; index cases were not sampled. Every row represents a bar, every square a serum. Sera are categorized as mpox-seropositive (red; positive for VACV-binding and MPXV-neutralizing antibodies) or mpox-seronegative (grey; negative for VACV-binding and MPXV-neutralizing antibodies). Sera testing positive for either VACV-binding or MPXV-neutralizing antibodies are shown as orange. (F) Percentage of households with seropositive members. (G) Serostatus from all individuals in households; index cases were not sampled. Every row represents a bar, every square a serum. Color-coding is identical to panel E. DRC, Democratic Republic of the Congo; VACV, vaccinia virus; MPXV, mpox virus.

Next, we developed a neutralization assay using an MPXV clade Ib isolate obtained from an mpox patient in DRC. To this end, we inoculated swab material on Vero cells, propagated the isolate to passage 2, and grew virus stocks on Calu-3 cells for neutralization assays (see **Methods** for details). The input material and all virus passages were sequenced to confirm virus identity and the absence of cell culture adaptations (GISAID accession number: EPI_ISL_19621388; **Figure S2**). Since MPXV clade Ib neutralization assays have not yet been described, we performed neutralization assays on 10 validation sera available at Erasmus MC (**Figure 1B**, red and grey circles). From these 10 control sera, 5 did not have VACV-binding antibodies (obtained from individuals known to be unvaccinated and unexposed) and 5 did (obtained from individuals known to have received historic smallpox and recent MVA-BN vaccination) (**Figure 1A**, red and grey circles). Indeed, the in-house developed neutralization assay with MPXV clade Ib demonstrated the presence of cross-neutralizing antibodies in the vaccinated individuals, but not the seronegative controls. Based on the negative control sera, we defined the cut-off as a titer threefold over background (>35).

We tested all available sera from the potentially exposed group and the pre-outbreak control group for the presence of clade Ib MPXV-neutralizing antibodies (**Figure 1B**). Positivity rates among the two bar groups, collected in November and December 2023, and May 2024, were 51% (61/120) and 42% (20/48), respectively. Among the sera collected from households, 44% (48/108) tested positive (**Table 2**). Using the combined presence of detectable binding antibodies by ELISA (>10) and confirmation by neutralization (>35) as the definition for mpox seropositivity, the overall seropositivities for the three groups were 18%, 17%, and 23%, respectively (**Table 2**). Similar to the VACV-binding antibodies, comparable MPXV-neutralizing antibody levels were detected among male and female individuals, and individuals from different age groups (**Figure S1B**).

While no clear correlation between VACV-binding and clade Ib MPXV-neutralizing antibody levels was observed, the samples from the different cohorts (seropositive exposed, pre-outbreak, and validation sera) clustered together (**Figure 1C**). Most of the individuals we defined as mpox-seropositive had high levels of MPXV-neutralizing antibodies, indicative of recent MPXV infection as described previously for MPXV clade IIb.^14^ The seropositive validation sera obtained from individuals with historic smallpox and recent MVA-BN vaccination had high levels of VACV-binding antibodies and intermediate levels of clade Ib MPXV-neutralizing antibodies (**Figure 1C**). Notably, many sera tested positive for the presence of MPXV-neutralizing antibodies in the absence of measurable titers by the VACV ELISA. This suggests that mpox seropositivity percentage we report here could be an underestimation of the actual seroprevalence.

As part of capacity building, we performed ELISA training at the University Teaching Hospital of Butare (CHUB; Huye, Rwanda). In total, 146 sera were tested at both sites (Figure S3) to allow comparison of local laboratory readiness for serological screening assays. Overall agreement between test results was 94% (137 out of 146). However, the testing at CHUB yielded 5 positives that could not be confirmed by ELISA at Erasmus MC, all of which did test positive in neutralization assays. Reversely, 4 samples tested positive at Erasmus MC but negative at CHUB, all of which were again neutralization-positive. This, combined with the detection of MPXV-neutralizing antibodies in the absence of VACV-binding antibodies, shows that further work is needed to do standalone serology based on ELISA.

To determine the role of PSW in MPXV clade Ib transmission in South Kivu, DRC, we then focused on the analysis of sera obtained from PSW and visitors of 25 bars (n=168) (**Tables 1** and **2**; **Figure 1D**). In 16 out of 25 bars (64%), there was serological evidence for MPXV circulation, defined as presence of both VACV-binding and MPXV-neutralizing antibodies in at least one serum sample obtained at that bar (**Figure 1E**). In 14 out of 16 bars with serological evidence for MPXV circulation, the serum sample(s) testing mpox-seropositive were obtained from PSW (88%). This serological evidence supports the hypothesis that sexual transmission is an important transmission route for MPXV clade Ib, which was earlier suggested by epidemiological and genomic surveillance of PSW.^9,10^ Especially in Kamituga, South Kivu, a mining region with a high number of PSW, this could have fueled the rapid spread of MPXV clade Ib.

Similarly, we used serological evidence to determine whether household transmission played a role in the rapid spread of MPXV clade Ib. Serum samples were obtained from 34 households with an index case within the household. The index cases themselves were not included in sampling. In 17 out of 34 households (50%), at least one serum sample obtained from contact cases tested mpox-seropositive, defined as both the presence of VACV-binding and MPXV-neutralizing antibodies (**Figure 1F**). In five of these households, there were at least two seropositive individuals (households 1, 4, 6, 20 and 43), of which four included at least one minor (<18 years; households 1, 4, 6 and 43) (**Figure 1G**). In five additional households (households 3, 7, 9, 18 and 19), we detected serological evidence of MPXV clade Ib infection in minors, often below the age of 10, suggesting that close-contact transmission via another route than sexual transmission had occurred.

Here, we show that sero-epidemiological studies are vital to understand the true extent of MPXV clade Ib circulation in the African region. The seropositivity rates in three different exposed groups ranged between 17% and 23%, using stringent definitions for seropositivity. When using the VACV-binding antibody ELISA as a screening assay, in combination with a newly developed MPXV clade Ib neutralization assay using a virus isolated directly from the field as confirmation, we show that individuals with detectable VACV-binding antibodies also had high neutralizing antibody levels. This is considered evidence of recent infection.^14^ High seroprevalence among PSW in bars confirmed that sexual transmission was a likely cause of the rapid MPXV clade Ib spread. Seropositivity in young children – as part of a study investigating follow-up of the introduction of mpox in a household – confirmed the occurrence of household transmission, likely via a close-contact non-sexual transmission route. As a next step, setting up local laboratories that perform serological assays is essential to monitor vaccine immunogenicity. Here, we used an ELISA based screening assay with confirmation by neutralization as evidence for seropositivity, but it remains to be determined whether the ELISA alone is sufficient for sero-epidemiological studies. Conclusively, serological studies could play a crucial role in determining vaccination policies and could guide decision makers towards relevant risk groups to be vaccinated to interrupt transmission.

## Acknowledgments

This study received partial funding from the EDCTP funded projects GREAT-LIFE (grant numbers 101103059, laboratory training) and JUA KIVU (grant number 101195116, study design and execution). Assay development at Erasmus MC was supported through the HERA project DURABLE (grant number 101102733) and the Netherlands Organization for Health Research and Development (ZonMw, grant agreement 10150022310035). We gratefully acknowledge the UK Medical Research Council (MRC) and the UK Foreign, Commonwealth & Development Office (FCDO) for their research support under the MRC-UKRI grant MC_PC_24001, which was awarded to local investigators, Kamituga Hospital, and the Division Provinciale de la Santé (DPS, Provincial Division of Health) in South Kivu, Bukavu. We also acknowledge the Wildlife Conservation Network (WCN) and Conservation Action Research Network (CARN) for the scholarship and research support they awarded to LMM. We would like to thank the DPS of South Kivu and Kamituga Health Zone for their collaboration during the study. Finally, the DPS acknowledges the support of the Bill & Melinda Gates Foundation [INV-078955 AIMS-NEI].

The following people from the GREAT-LIFE mpox group provided technical assistance to the study: Nadine Bubala Malyamungu, Felix Camarade Habarugira, Francisco Bacon Benimana, Chantal Umubyeyi and Leandre Mutimbwa Mambo. We acknowledge Martin E. van Royen (Department of Pathology, Erasmus University Medical Center, Rotterdam, the Netherlands) for technical expertise regarding imaging, Eric van Gorp (Department of Viroscience, Erasmus University Medical Center, Rotterdam, the Netherlands) for the collection of validation sera in the COVA study, and Sandra Scherbeijn (Department of Viroscience, Erasmus University Medical Center, Rotterdam, the Netherlands) for performing HIV serology. Finally, we greatly thank Christian Gortázar (SaBio Instituto de Investigación en Recursos Cinegéticos, Universidad de Castilla-La Mancha y CSIC, Ciudad Real, Spain) and Trudie Lang (The Global Health Network (TGHN), University of Oxford, Oxford, United Kingdom) for their advice.

## Data availability

Data from the present study are not part of public databases but are available upon reasonable request from the corresponding author. Patient-related data may be subject to patient confidentiality, or unavailable due to the use of pseudonymized data. This study used unique materials, which were custom-made for specific analyses (ELISA antigens and virus stocks). Materials are available upon reasonable request, will be released via a material transfer agreement and can otherwise be obtained via the included experimental protocols in the Methods. The consensus sequence of the clade Ib MPXV isolate is available on GISAID under the accession number EPI_ISL_19621388.

## Code availability

Code and software use has been disclosed where applicable. GitHub repositories used for MPXV sequencing are included in the respective section of the Methods.

## Author contributions

LMM, LMZ, PN, JCU, SO, FA, JPM, JBM, JMN, FSB, MPGK, and RDDV conceptualized the study. LMM, PN, JCU, JPM, JBM, and FSB were involved in sample collection. LMM, LMZ, LS, BEV, BBOM, CHGvK, and RDDV contributed to data acquisition and investigation. LMZ, LS, BBOM and RDDV performed the formal analysis. LMZ, PN, JCU, FA, MPGK, and RDdV acquired funding. LMM, LMZ, PN, JCU, MPGK, and RDDV performed project administration. PN, JCU, FSB, MPGK, and RDDV supervised the study. RDDV and MPGK wrote the original draft of the manuscript. All authors reviewed, edited and approved of the final version of the manuscript.

## Methods

### Ethics statement

For the samples obtained in DRC, ethical clearance was obtained from the ethical review committee of the Catholic University of Bukavu (UCB/CIES/NC/022/2023) and a tripartite Memorandum of Understanding signed between the Provincial Health Division (Division Provinciale de la Santé, DPS), Stansile and the Erasmus MC. All participants provided informed consent. In case the participant was a minor, parental consent was obtained. The 10 validation sera were collected from laboratory workers in the Netherlands who received MVA-BN vaccination for safety reasons as employees of a Biosafety Level 3 (BSL-3) laboratory under the Erasmus Medical Center vaccination cohort (COVA) biobanking study protocol (MEC-2014-398). The protocol was approved by the Erasmus MC Medical Ethics Review Committee. Written informed consent was obtained from all study participants.

### Participants

Studies were performed in two cohorts: a potentially exposed cohort and a pre-outbreak control cohort. Additional validation samples were included.

#### Potentially exposed bar cohorts

For this part of the study, serum samples were collected in November and December 2023 (n=120), and May 2024 (n=48) from bars that reported at least one confirmed, suspected, or probable mpox case in two rounds. Index cases were not included in this study. The amount of contact between index cases and sampled participants in bars is not known. None of the participants were symptomatic at the time of sampling. Demographic data from all participants was collected. A confirmed mpox case was defined as an individual with a laboratory-confirmed MPXV infection as tested by the National Institute for Biomedical for Research (INRB). A suspected mpox case was defined as an individual with acute illness who had fever, intense headache, myalgia, and back pain, followed by one to three days of a progressively developing rash, often starting on the face and spreading to the rest of the body. A probable mpox case was defined by an individual meeting the clinical definitions described for suspected cases, who had an epidemiological link to a confirmed or suspected case but was not laboratory-confirmed.

#### Potentially exposed household cohort

In addition to samples collected in bars, we performed contact tracing for confirmed mpox cases with a link to a household and collected samples from the family members (contact cases). A contact case was defined to have had contact (non-physical, physical, sexual, respiratory, fomites, or any other form) with the confirmed mpox case. The respective index cases were not part of this study. None of the participants were symptomatic at the time of sampling. Household samples were collected between September 2023 and May 2024 (n=108). Demographic data from all participants was collected.

#### Pre-outbreak cohort and validation sera

For the pre-outbreak cohort, randomly selected archived serum samples collected at least two years prior to the ongoing MPXV clade Ib outbreak, in an area that has not reported mpox before, were used (n=51). Samples were collected in a hospital with an outpatient clinic that, among other things, distributes anti-retroviral therapy for HIV. For the validation sera, 10 sera available at Erasmus MC from laboratory workers who received MVA-BN vaccination for safety reasons as employees of a Biosafety Level 3 (BSL-3) laboratory were used. Of these 10 sera, 5 were used as negative samples (obtained from individuals known to be unexposed and before MVA-BN vaccination) and 5 were used as positive samples (obtained from individuals known to have received historic smallpox and recent MVA-BN vaccination).

### Sampling

Five ml of blood was collected in dry tubes from which 2 ml of serum was obtained by centrifugation, and subsequently stored at −20 ºC before assays were performed. To perform virus isolations, lesion swabs from the genital area of suspected mpox cases were collected between 12 August and 3 September 2024, and stored in virus transport medium at −20 ºC before shipment on dry ice to Erasmus MC.

### Isolation and propagation of MPXV clade Ib

MPXV clade Ib was isolated from swab material obtained from an mpox patient in DRC. Presence of MPXV clade Ib was confirmed by real-time PCR (RT-PCR) and sequencing.^16^ The virus transport medium in which the swab was stored was inoculated on Vero cells in Advanced DMEM/F-12 (Gibco), supplemented with 10 mM HEPES (Gibco), 1×GlutaMAX (Gibco), and 1×Primocin (InvivoGen), referred to as AdDF+++, and the isolated virus was propagated to passage 2 on Vero cells. Each passage was harvested upon development of cytopathic effect across most of the monolayer using freeze-thaw cycles to release the intracellular mature virions (IMV) from the cells.^14^ Virus stocks for neutralization assays were grown on Calu-3 cells. To this end, Calu-3 cells were inoculated at a multiplicity of infection of 0.05 in AdDF+++ and harvested as described above. All experimental work involving infectious MPXV were performed in a class II biosafety cabinet under BSL-3 conditions. MPXV stock titers were determined as described previously.^14^

### MPXV sequencing

Original input material and all virus passages were submitted to whole genome sequencing to confirm virus identity and exclude cell culture adaptations. DNA was randomly amplified using sequence-independent single-primer amplification.^17^ Sequencing libraries were generated using the Native Barcoding Kit 24 V14 (Oxford Nanopore Technologies), sequenced on a PromethION flow cell (R10.4.1) with super-accurate basecalling using Dorado v7.4.12 (ONT). MPXV consensus was generated using Virconsens (https://github.com/dnieuw/Virconsens) with >30x coverage cut-off and uploaded to GISAID (EPI_ISL_19621388). The consensus was aligned with 163 publicly available MPXV clade Ib sequences from GISAID (Table S1) using squirrel v1.0.12 (https://github.com/aineniamh/squirrel) followed by phylogenetic analysis using IQ-TREE v2 (https://github.com/iqtree/iqtree2) (Figure S2). Clade assignment and quality checks were performed on Nextclade v3.9.1 (https://github.com/nextstrain/nextclade).

### Detection of VACV-binding IgG antibodies

Binding IgG antibody levels against VACV were measured both at Erasmus MC as well as on site during a workshop at the CHUB, Huye, Rwanda, using an in-house developed ELISA as described previously.^14^ Identical antigens were used. Briefly, high-binding 96-well ELISA plates (Corning) were coated with the cell lysate of either VACV-infected or mock-treated HeLa cells diluted 1:250 in PBS for 1 h at 37 ºC. After coating, plates were washed with a washing buffer (PBS + 0.05% Tween20 [Merck]) and blocked for 1 h at 37 ºC using a blocking buffer (2% skim milk powder [Merck] in washing buffer). Fivefold dilution series of the heat-inactivated sera (30 min, 56 ºC) in blocking buffer starting at a 1:10 dilution were prepared, added to the plates, and incubated overnight at 4 ºC. Afterwards, plates were washed with a washing buffer and incubated for 1 h at 37 ºC with horseradish peroxidase (HRP)-conjugated rabbit anti-human IgG (1:6,000; Dako) diluted in blocking buffer. After another washing step, plates were developed with TMB peroxidase substrate (SeraCare). Absorbance was measured at 450 nm using a microtiter plate reader (Erasmus MC: Anthos 2001 microplate reader; CHUB: BioTek 800 TS) and corrected by subtracting absorbance at 620 nm. A net OD450 response was determined by subtracting the OD450 value from plates coated with mock-treated cell lysate from those coated with VACV-infected cell lysate. A positive reference serum pool was included on every plate and used to calculate 30% endpoint titers by transforming the net OD450 responses per sample to this positive control S-curve.

### Detection of MPXV clade Ib-neutralizing antibodies

A plaque reduction neutralization test (PRNT) using the newly isolated MPXV clade Ib was developed based on the MPXV clade IIb neutralization assay that we previously described.^14^ Essentially, Vero cells were seeded 1 day prior in 96-well cell culture plates. Heat-inactivated sera (30 min, 56 ºC) were twofold serially diluted in AdDF+++ before adding an equal volume of medium containing 300 PFU MPXV clade Ib. All measurement were performed in duplicate. The virus-serum mix was incubated for 1 h at 37 ºC before being transferred to the cell culture plates. After incubation for 24 h at 37 ºC and 5% CO2, the cells were fixed with 4% paraformaldehyde for 30 min and permeabilized with 70% ethanol. Following washing with PBS and blocking in 0.6% BSA and 0.1% Triton X-100 in PBS for 30□minutes, MPXV clade Ib was visualized by overnight staining with rabbit-anti-VACV-FITC (1:1,000 in blocking buffer; Abbexa) at room temperature. Cell nuclei were counterstained with Hoechst33342 (Thermo Fisher Scientific). Imaging was done using the Opera Phenix spinning disk confocal HCS system (PerkinElmer) equipped with a ×10 air objective (NA 0.3) and 405-nm and 488-nm solid-state lasers. Infected cells were quantified using the Harmony software (version 4.9, PerkinElmer). The dilution that would yield 50% reduction of plaques (PRNT50) compared with the infection control was estimated by determining the proportionate distance between two dilutions from which an endpoint titer was calculated. When no neutralization was measured, the PRNT50 was assigned a value of 10. The cut-off for positivity was defined as three times the background of a set of five negative validation sera (≤35, negative; >35, positive).

### HIV serological analysis

Initial testing for the presence of HIV-specific antibodies was done on-site in 178 out of the 327 serum samples using the Determine™ HIV Early Detect kit (Abbott). Additionally, all 327 serum samples were screened for the presence of HIV-specific IgG antibodies at Erasmus MC to validate and complete the dataset using a diagnostic combination of a Liaison XL HIV Ab/Ag immunoassay (DiaSorin, Italy) and a line immunoassay (Inno-Lia HIV I/II; FujiRebio) on an Auto-LiPA (FujiRebio) automated system.^18^ All assays were performed according to the manufacturer’s instructions. Sera above the cut-off for positivity (S/CO□≥□1) were assessed by a line immunoassay to confirm the presence of HIV-specific antibodies. Sera with a S/CO□<□1 were considered non-reactive. Samples were considered seropositive if they tested positive in the screening assay and presence of HIV-specific antibodies was confirmed by a secondary method (detection of reactivity either using the line immunoassay and/or during on-site testing). Samples were considered seronegative if they tested negative in the screening assay. Out of the total of 327 serum samples, 15 yielded an inconclusive test result and four could not be tested due to insufficient sample volumes.

### Statistical analysis

A descriptive analysis was performed without statistical comparisons. Simple linear regression was performed on log-transformed VACV ELISA titers to determine the correlation between the ELISA performed at Erasmus MC (Netherlands) and CHUB (Rwanda) by Spearman’s r.

