## Supplementary Material for "Serological evidence supports the transmission of clade Ib mpox virus by professional sex workers and spread within households in South Kivu, DRC"

**Supplementary tables**

**Table S1.** Data acknowledgement table with GISAID EPI numbers of MPXV clade Ib sequences used for phylogenetic analysis.

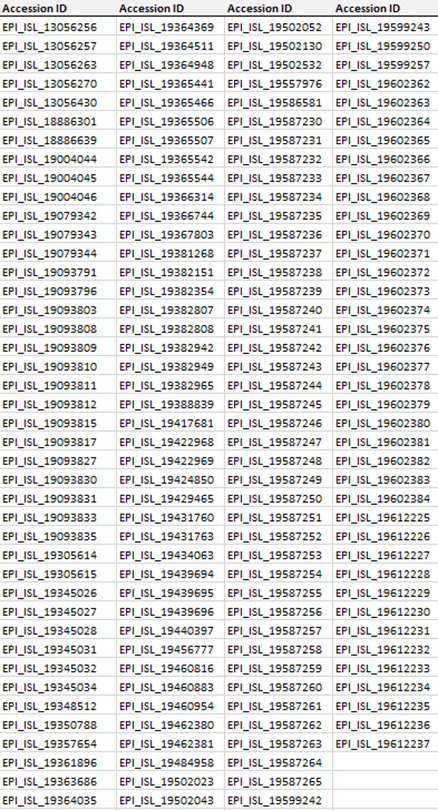

**Supplementary figures**

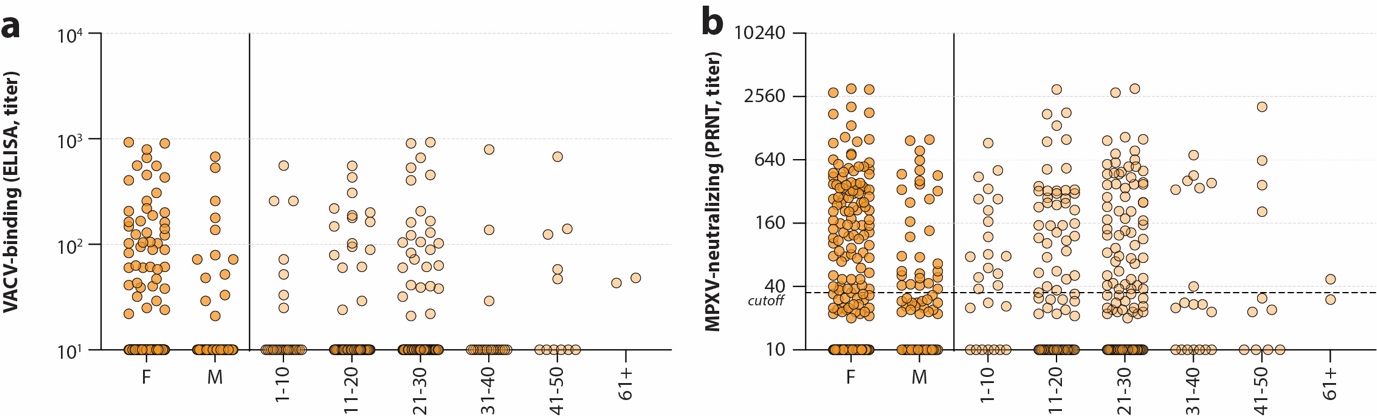

**Figure S1. VACV-binding and MPXV-neutralizing antibodies stratified by biological sex and age group.** (A) VACV-binding antibodies measured in sera obtained from bars and households (**Figure 1A**) stratified by biological sex (M: male; F: female) and age group. (B) MPXV-neutralizing antibodies measured in sera obtained from bars and households (**Figure 1B**) stratified by biological sex and age group.

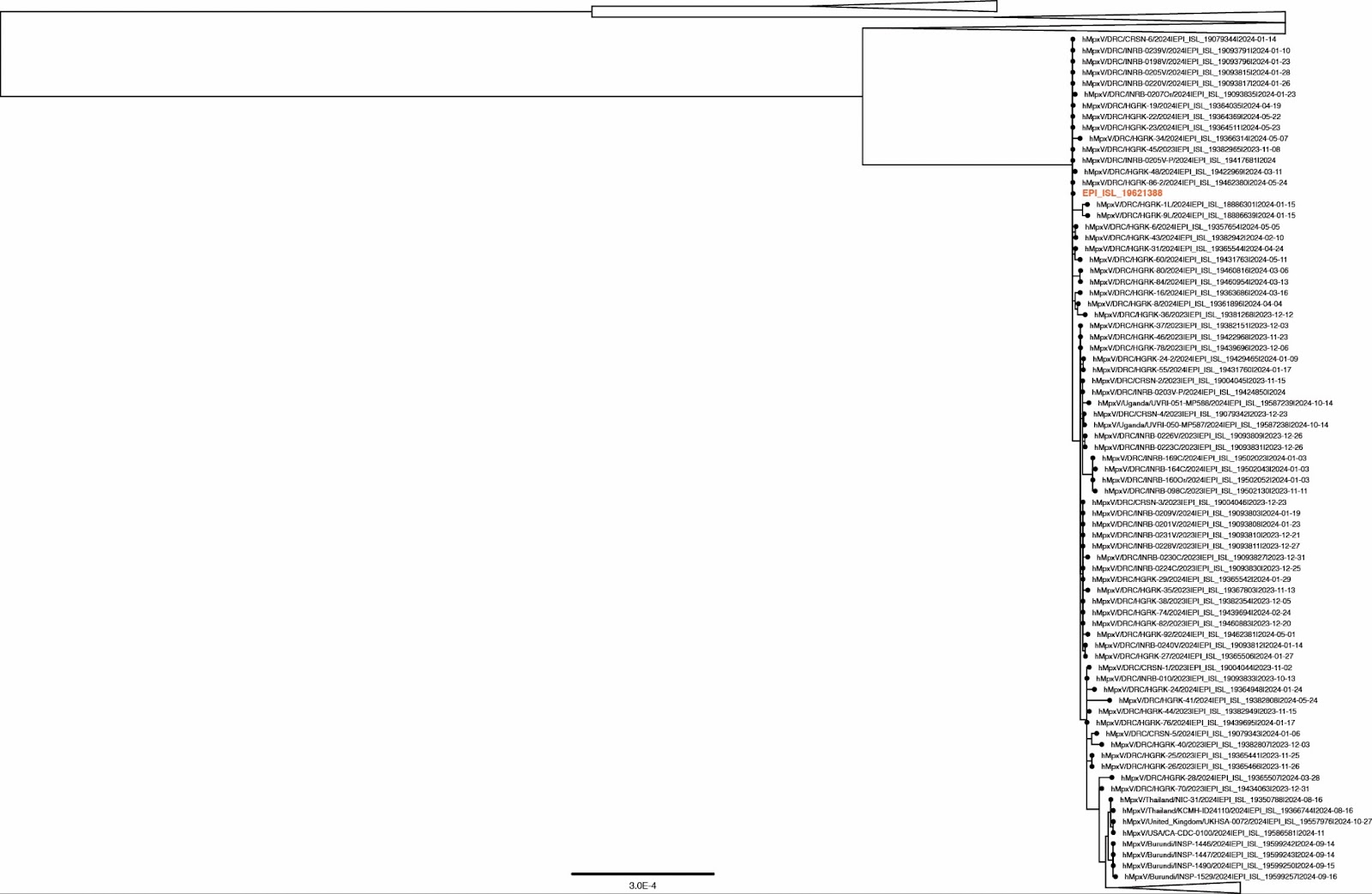

**Figure S2.** **Phylogenetic analysis of the generated MPXV sequence (highlighted in orange) with publicly available MPXV clade Ib sequences**. The collapsed section at the bottom includes a large Ugandan sequence cluster (n=61) and cases from Sweden (n=2), Kenya (n=1), Germany (n=1), India (n=1) and the UK (n=1). Original swab material consensus sequence has 10,654x depth, 100% genome coverage and was classified as MPXV clade Ib.

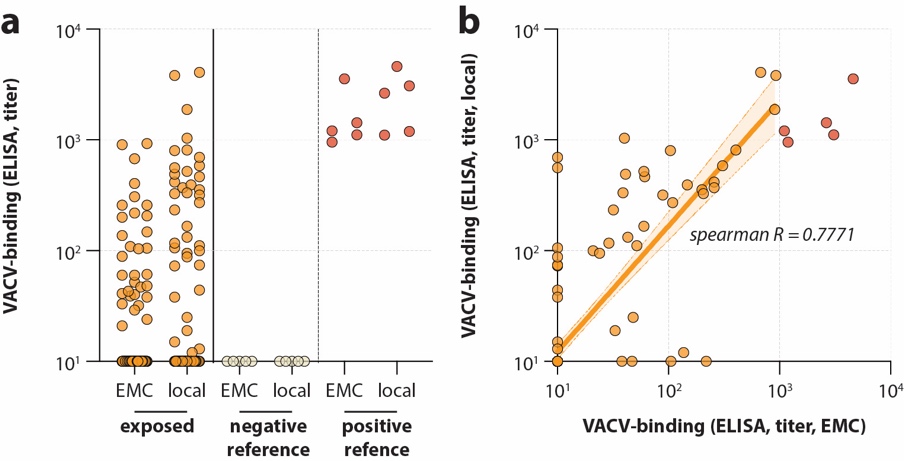

**Figure S3. VACV-binding antibodies measured at Erasmus MC or a local laboratory.** (A) Antibodies were measured using an in-house ELISA in exposed individuals (orange) and n=10 validation sera (grey and red). A subset of samples was measured at both Erasmus University Medical Center (EMC), Rotterdam, the Netherlands, and during a workshop at a local laboratory at the University Teaching Hospital of Butare (CHUB), Huye, Rwanda. (B) Correlation between VACV-binding antibody titers measured by ELISA performed at EMC and CHUB of samples from exposed individuals (orange) and validation samples (red). Line shows linear regression on log-transformed titers.
